# No Car, no Care? Assessing Transportation-Related Barriers to Specialized Oncology in Rural Germany

**DOI:** 10.1101/2025.11.26.25339657

**Authors:** Jonas Kapitza, Laura Bernhardt, Florian Lüke, Anja Sedlmeier, Tatjana Einwag, Julia Maurer, Gerhard Schenkirsch, Jutta Zeihsel, Markus Metzler, Alexander Kerscher, Markus Krebs

## Abstract

**Background:** Access to precision oncology not only depends on the provision of medical services but also on the practical ability of patients to reach their cancer center. Transportation barriers may substantially limit this access, particularly for patients living in rural areas.

**Methods:** We analyzed travel times to the four sites of the German WERA cancer center network (Würzburg, Erlangen, Regensburg, and Augsburg) for private car use and public transportation. To assess clinical relevance, we calculated travel times for a real-world cohort of WERA cancer patients from 2024 (n=63,524). Taxi accessibility was evaluated for given transportation budgets.

**Results:** Within a 2-hour travel time, private cars provided access to our cancer centers from nearly all areas of Bavaria and several neighboring regions, connecting 98.1% of our patients to WERA. In contrast, public transportation provided access to at best 74.5% of patients (2-hour travel time). Moreover, public transportation accessibility was highly time-dependent – with connectivity rates dropping sharply in the evening, particularly in rural areas. Finally, our analysis of taxi services showed that, for a round-trip budget of €100, 45.9% of our patient cohort had access to their closest WERA site. For virtual budgets of €200 and €300, accessibility rates rose to 71% and 87.5%, respectively.

**Conclusions:** Public transportation to our cancer centers is limited, especially during evening and nighttime hours. Switching to taxi services may bridge accessibility gaps but often entails a financial burden for patients and the healthcare system. Future outreach measures should “mind the gap” of public transportation.

**Implications for Practice:** Receiving care from a tertiary cancer center often requires regular site visits. Patients from distant and rural areas face considerable travel burdens, particularly when they lack access to a private car. Measures aimed to improve patient access should consider the limited accessibility of our centers via public transportation – especially during nighttime.

## INTRODUCTION

Ensuring fair and equal access to state-of-the-art cancer care proves challenging for healthcare providers and authorities worldwide, with patients living in rural regions being particularly at risk. Multimodal treatment concepts and participation in clinical trials frequently require repeated visits to specialized cancer centers, which impose substantial time, organizational, and financial burdens, especially for patients residing in remote and rural areas.^1–4^

Previous studies indicate that mobility-related barriers can cause treatment delays and compromise adherence to cancer therapies.^5^ A high travel burden has consequently been associated with inferior treatment outcomes in several studies. For Germany, registry-based analyses have shown a worse prognosis for patients with head and neck cancer, breast cancer and melanoma when living in rural areas.^6–7^ In the United States, greater travel distances to radiation therapy facilities have been associated with higher mastectomy rates, less use of adjuvant radiotherapy, and poorer survival among women with breast cancer.^8–10^ Similarly, studies from the United Kingdom have reported higher tumor stages at diagnosis and less favorable outcomes across multiple malignancies in regions with restricted access to specialized centers.^11–12^ Given this evidence, it is crucial to examine regional differences in access to specialized oncology care.

In Germany, the WERA alliance, consisting of the four comprehensive cancer centers in Würzburg, Erlangen, Regensburg, and Augsburg, provides cancer care for patients from its mainly rural catchment area.^13–14^ We therefore analyzed the accessibility of WERA’s cancer centers by different modes of transportation – private cars, public transportation and taxi services. Moreover, we investigated the public transportation accessibility depending on the time of day and the potential financial costs of using taxi services as the remaining mode of transportation. To assess the practical relevance of our findings, we calculated the traveling times for a real-world cohort of patients receiving cancer care by the WERA alliance.

We are convinced that a better understanding of transportation barriers will improve patient access to precision oncology – ensuring that all patients, irrespective of their place of residence or their financial situation, receive the best possible care. Our findings could support targeted policies and outreach programs aimed at reducing transportation-related challenges.

## MATERIALS AND METHODS

### Study design and model setup

To assess the spatial and temporal accessibility of the four WERA cancer centers, we generated isochrones using a transport model in the open-source geographic information system QGIS 3.34.^15^ The analysis covered three modes of transport: private car, public transportation and taxi services. For private car and public transportation, we focused on accessibility in terms of time. For using taxi services, we calculated resulting financial costs. Accessibility intervals for private cars and public transportation were set at ≤30, ≤60, ≤90 and ≤120 minutes. To reflect the temporal accessibility of the WERA sites and to account for day/night variations, we performed public transportation calculations for each hour over a 24-hour period (with Wednesday, August 20^th^, 2025, serving as the reference date). The public transportation model applied the following conditions:

- Direction of travel headed towards the closest WERA site.
- Travelling started at the beginning of each hour (e.g., 01:00, 02:00).
- A maximum walking time of 15 minutes was allowed from the starting point to the departure station and from the arrival station to the cancer center.
- If a transfer required walking (e.g., between train platforms), a maximum walking time of 10 minutes was factored in.
- All walking times were included in the total travel time.

To assess taxi transportation and its associated costs, we contacted 20 local taxi companies via email five companies responded. Based on these responses and the official taxi fares of the cities of Würzburg^16^, Erlangen^17^, Regensburg^18^ and Augsburg^19^, we calculated costs for one-way and round trips. As most companies reported a maximum waiting time of 15 minutes at the respective cancer center, we excluded waiting time costs from further calculations.

### Patient cohort

We further assessed the practical relevance of our findings and calculated traveling times for cancer patients treated at one of the four WERA sites in 2024. For this study, the medical history of patients had to contain at least one of the following events:

- First diagnosis of a malignancy by a WERA site in 2024.
- Cancer treatment as a WERA in-house patient in 2024.
- Tumor board discussion by a WERA site in 2024 (in-house patients and external patients receiving a second opinion).
- Change in the course of cancer disease (e.g., partial response, PR) in 2024 and subsequent reporting to the state cancer registry.

Selected patients were required to have a known postal code and residence in Germany. Altogether, following our search algorithm resulted in a final cohort of 63,524 WERA patients distributed over 3,474 German postal code areas (Table 1, Figure 1A).

**Table 1.** Distribution of cancer patients and postal code areas across the sites of the WERA cancer center network.

| WERA Site | Patients (n) | Number of Postal Code Areas |
| --- | --- | --- |
| Würzburg | 16,542 | 1862 |
| Erlangen | 20,241 | 1563 |
| Regensburg | 12,727 | 964 |
| Augsburg | 14,014 | 866 |

**Figure 1.**
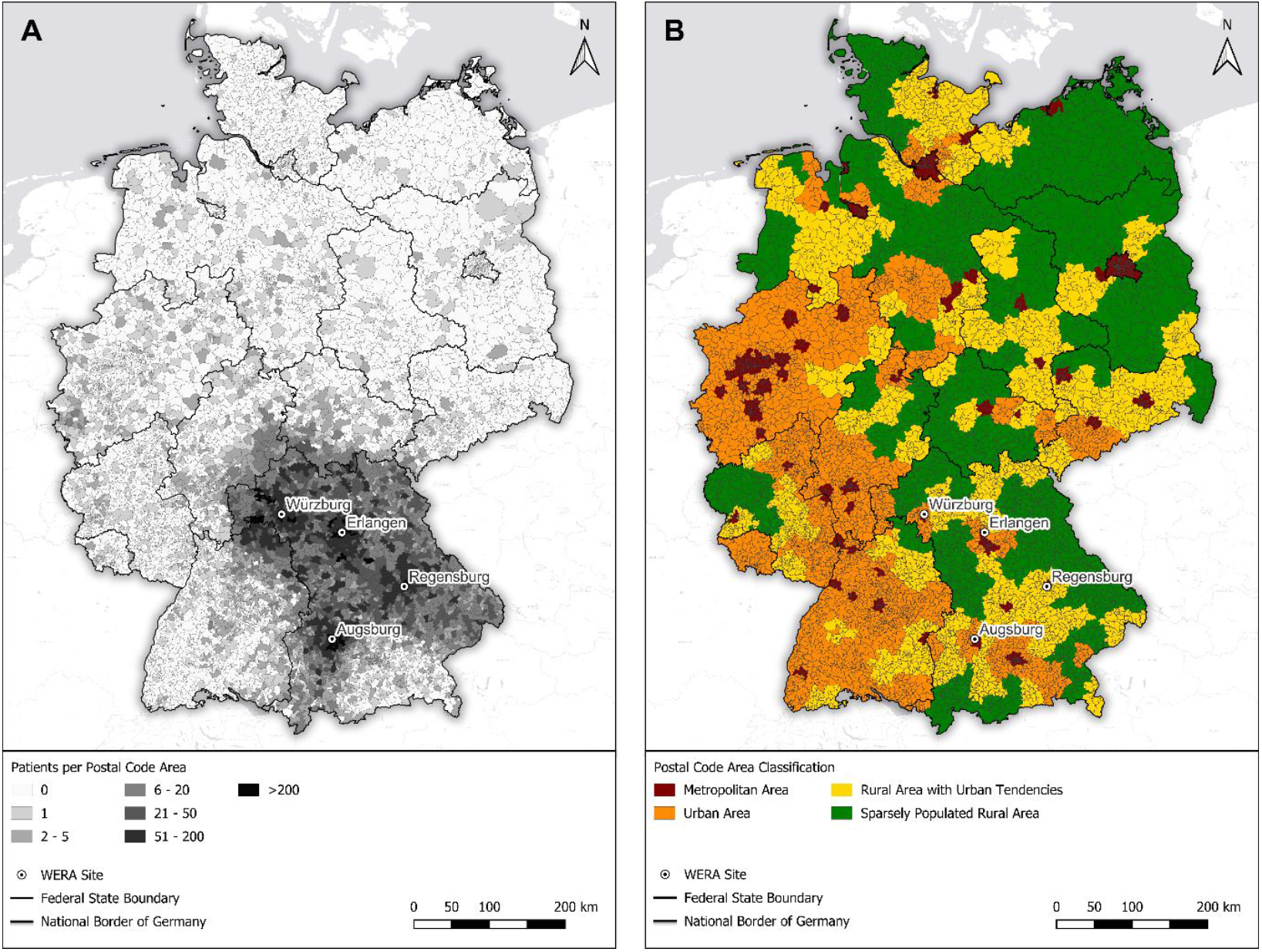
**(A)** Nationwide distribution of the places of residence of WERA patients by postal code area, based on data from 2024 (n=63,524). **(B)** Nationwide classification of postal code areas using spatial categories: Metropolitan areas (dark red), urban areas (orange), rural areas with urban tendencies (yellow), and sparsely populated rural areas (green).

Exclusively using retrospective and anonymous data, our study was conducted in accordance with the European General Data Protection Regulation (GDPR, Art. 9),^20^ the German Federal Data Protection Act (BDSG, §22),^21^ and the Bavarian Hospital Act (BayKrG, Art. 27).^22^ In addition, the ethics committee of the University of Regensburg approved our study (25-4415-180).

### Geospatial classification

To classify the postal code areas of our catchment area in terms of urbanity and rurality, we first assigned them to districts, since spatial classifications in Germany are exclusively provided at the district level. The assignment of the postal code areas to districts relied on publicly available data from Opendatasoft.^23^ Subsequently, districts were assigned to spatial classifications provided by the Federal Institute for Research on Building, Urban Affairs and Spatial Development (BBSR).^24^ Figure 1B shows the classification of all German postal code areas into four categories – “metropolitan” (dark red), “urban” (orange), “rural with urban tendencies” (yellow), and “sparsely populated rural areas” (green).

Finally, we generated isochrones for different modes of transportation and times of the day to delineate areas from which a WERA site could be reached within predefined travel time intervals. A postal code area was considered accessible if it was intersected by the corresponding isochrone. Based on this classification, we calculated accessibility rates as the proportion of patients residing in postal code areas that allowed timely access to a WERA site.

## RESULTS

### Accessibility of the WERA sites by private car

First, we calculated the spatial accessibility of the WERA sites by private car, serving as a benchmark for comparison with other modes of transport. Apart from some southern areas in the foothills of the Alps, WERA sites were accessible throughout Bavaria within a maximum travel time of 120 minutes (Figure 2, yellow). Additionally, parts of southern Thuringia, southeastern Hesse, and large areas of eastern Baden-Württemberg were within this 120-minute range. In fact, most Bavarian postal code areas had travel times of 90 minutes or less to the closest WERA site (Figure 2, green).

**Figure 2.**
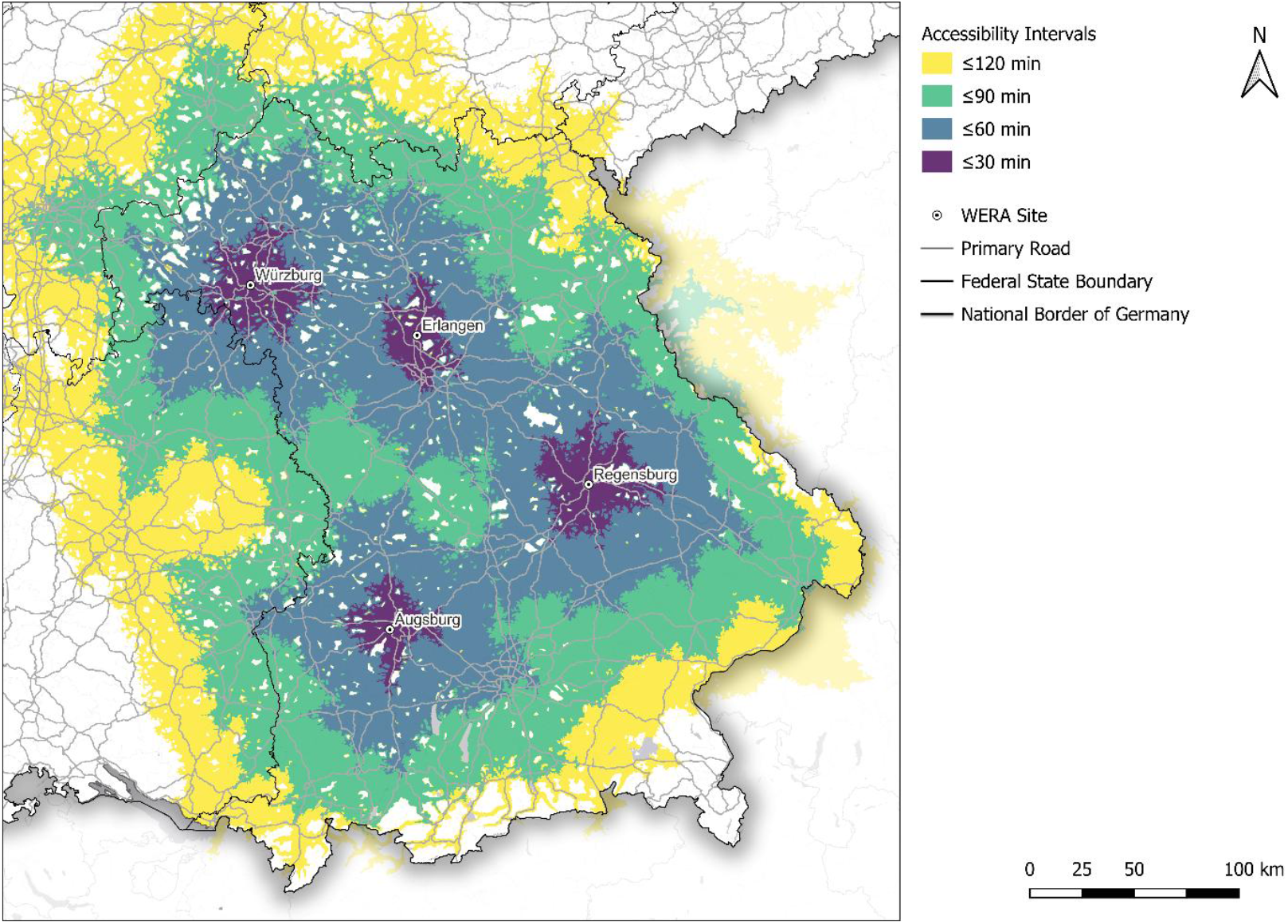
Accessibility of the four WERA sites by private car within given time intervals: ≤30 minutes (purple), ≤60 minutes (blue), ≤90 minutes (green), and ≤120 minutes (yellow). Uncolored areas either did not fall within the 120-minute accessibility range or failed to meet basic model requirements, such as being within a 15-minute walk of a road accessible to both pedestrians and cars. These areas included forests, large bodies of water, mountain ranges, and regions designated for military or environmental purposes.

### Public Transport Accessibility of WERA sites

Since not all potential patients have access to a private car, we analyzed the spatial and temporal accessibility of the WERA sites via public transportation. At first sight, public transportation covered a significantly smaller area within given timeframes. In contrast to private car use, public transportation accessibility highly depends on the time of traveling. Figure 3 depicts the accessibility at four exemplary time points: 12:00, 17:00, 21:00, and 01:00. In general, public transportation accessibility depended on existing rail and bus networks, with the highest accessibilities observed in urban regions in or close to Würzburg, Erlangen, Regensburg, and Augsburg. Regions and cities such as Greater Munich (southeast of Augsburg), Nuremberg (south of Erlangen), and Fulda (north of Würzburg), also had potential access to WERA sites within 120 minutes during daytime (Figure 3A, B).

**Figure 3.**
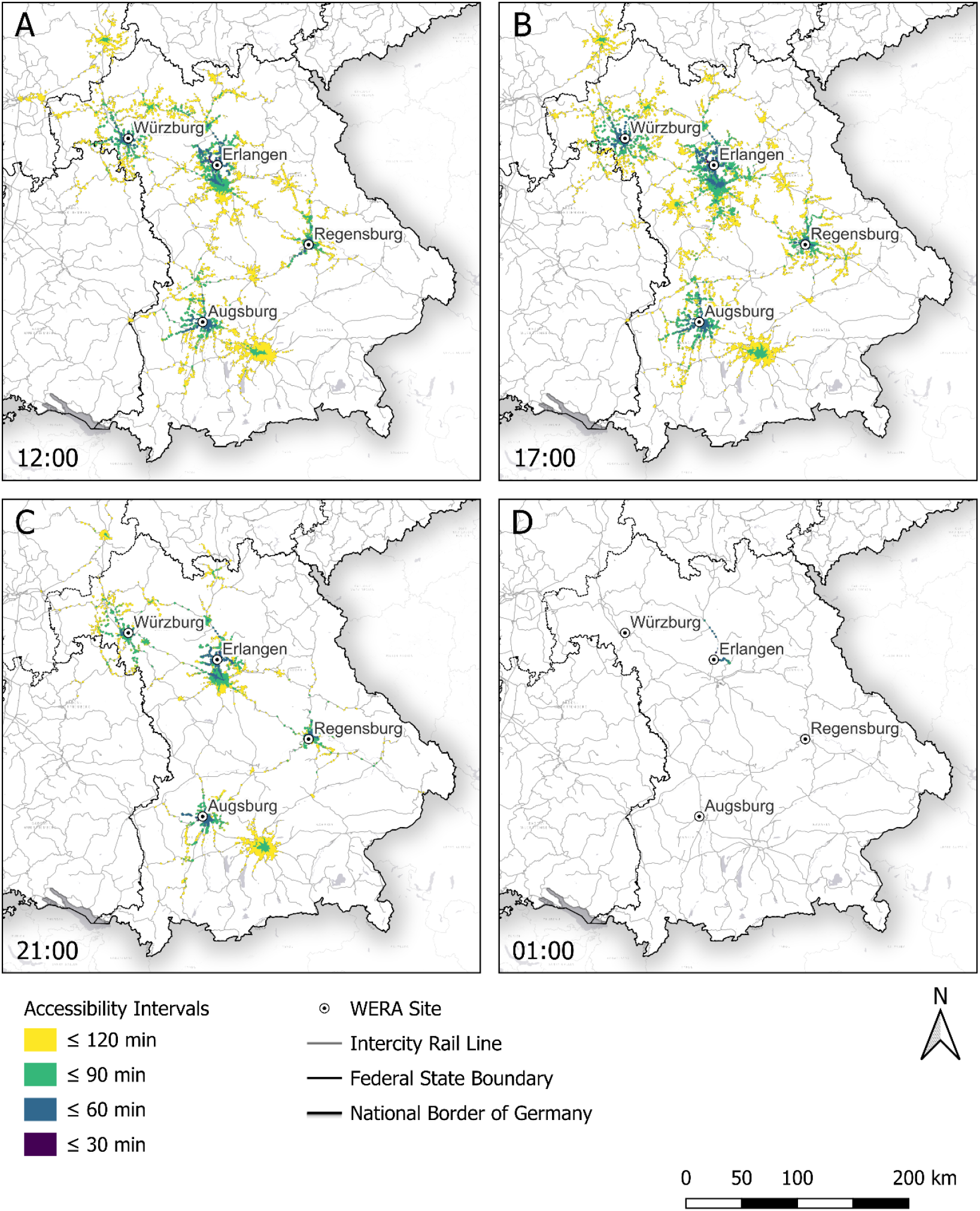
Accessibility of the four WERA sites by public transportation within predefined time intervals: ≤30 minutes (purple), ≤60 minutes (blue), ≤90 minutes (green), and ≤120 minutes (yellow), depending on the time of day: **(A)** 12:00, **(B)** 17:00, **(C)** 21:00, and **(D)** 1:00.

Of note, public transportation covered a significantly smaller area compared to private cars, with a maximum coverage of 7,850 km^2^ for public transportation at 17:00 (Figure 3B and Figure S1) compared to 105,900 km^2^ for a 120-minute range using a private car. In the evening and night hours, public transportation accessibility dramatically diminished. By 21:00 (Figure 3C), many regions that were connected during the day fell outside the 120-minute travel range – resulting in a sharply dropped coverage of 3,570 km^2^ (Figure S1). By 01:00 (Figure 3D), the remaining few bus and train connections reaching WERA within 120 minutes were solely located in the Erlangen-Nuremberg metropolitan area, covering only 71 km^2^ (Figure S1).

### Real-world accessibility for WERA patients

To assess the actual impact of limited public transportation accessibility, we calculated travel times for a real-world cohort of n=63,524 patients treated by the WERA alliance in 2024 (Figure 1A). We therefore assigned our patients’ places of residence to one of four official categories – ranging from “metropolitan areas” to “sparsely populated rural areas” (Figure 1B). As shown in Table 2, 20.5% of our patients came from “metropolitan areas” characterized by cities with at least 100,000 inhabitants. Another 22.5% of patients lived in “urban areas”. Of note, more than half of our study population (57.0%) lived in areas categorized as more rural, with 30.2% originating from “rural areas with urban tendencies” and 26.8% living in “sparsely populated rural areas”. These results underscore the relevance of accessibility from rural areas for the WERA cancer centers.

**Table 2.** Distribution of WERA cancer patients (n=63,524) by spatial category of place of residence. Postal code areas were assigned according to the official classification of the Federal Institute for Research on Building, Urban Affairs and Spatial Development (BBSR).

| Spatial category | Characteristics of each category | Number and share of WERA patients per category |  |
| --- | --- | --- | --- |
|  |  | n | % |
| Metropolitan area<br>(*Kreisfreie Großstadt*) | Cities with $\geq 100,000$ inhabitants | 13,008 | **20.5** |
| Urban area<br>(*Städtischer Kreis*) | $\geq 50\%$ population in large/medium cities and population density $\geq 150$ inhabitants/km<sup>2</sup> | 14,309 | **22.5** |
| Rural area with urban tendencies<br>(*Ländlicher Kreis mit Verdichtungsansätzen*) | $\geq 50\%$ population in large/medium cities, but population density $< 150$ inhabitants/km<sup>2</sup> or $< 50\%$ population in large/medium cities, but population density $\geq 100$ inhabitants/km<sup>2</sup> | 19,159 | **30.2** |
| Sparsely populated rural area<br>(*Dünn besiedelter ländlicher Kreis*) | $< 50\%$ population in large/medium cities and population density $< 100$ inhabitants/km<sup>2</sup> | 17,048 | **26.8** |

We next calculated travel times via private car and public transportation for all postal code areas harboring patients of our study cohort. In line with our previous results (Figure 2), a high proportion of our study cohort had access to WERA via private car – with 55.0% of patients reaching the closest WERA site within 30 minutes, 86.2% within 60 minutes, 96.4% within 90 minutes, and 98.1% within 120 minutes.

Compared to private car use, accessibility rates by public transportation were generally lower and varied by the time of day and place of residence. During daytime (between 05:00 and 21:00), approximately 15% of patients reached a WERA site within 30 minutes, around 36% within 60 minutes, and about 75% had access to WERA within 120 minutes (Figure 4A). During the middle of the night (between 01:00 and 03:00), public transportation accessibility dropped sharply – with a maximum patient share of 13.3% reaching a WERA site within 120 minutes.

**Figure 4.**
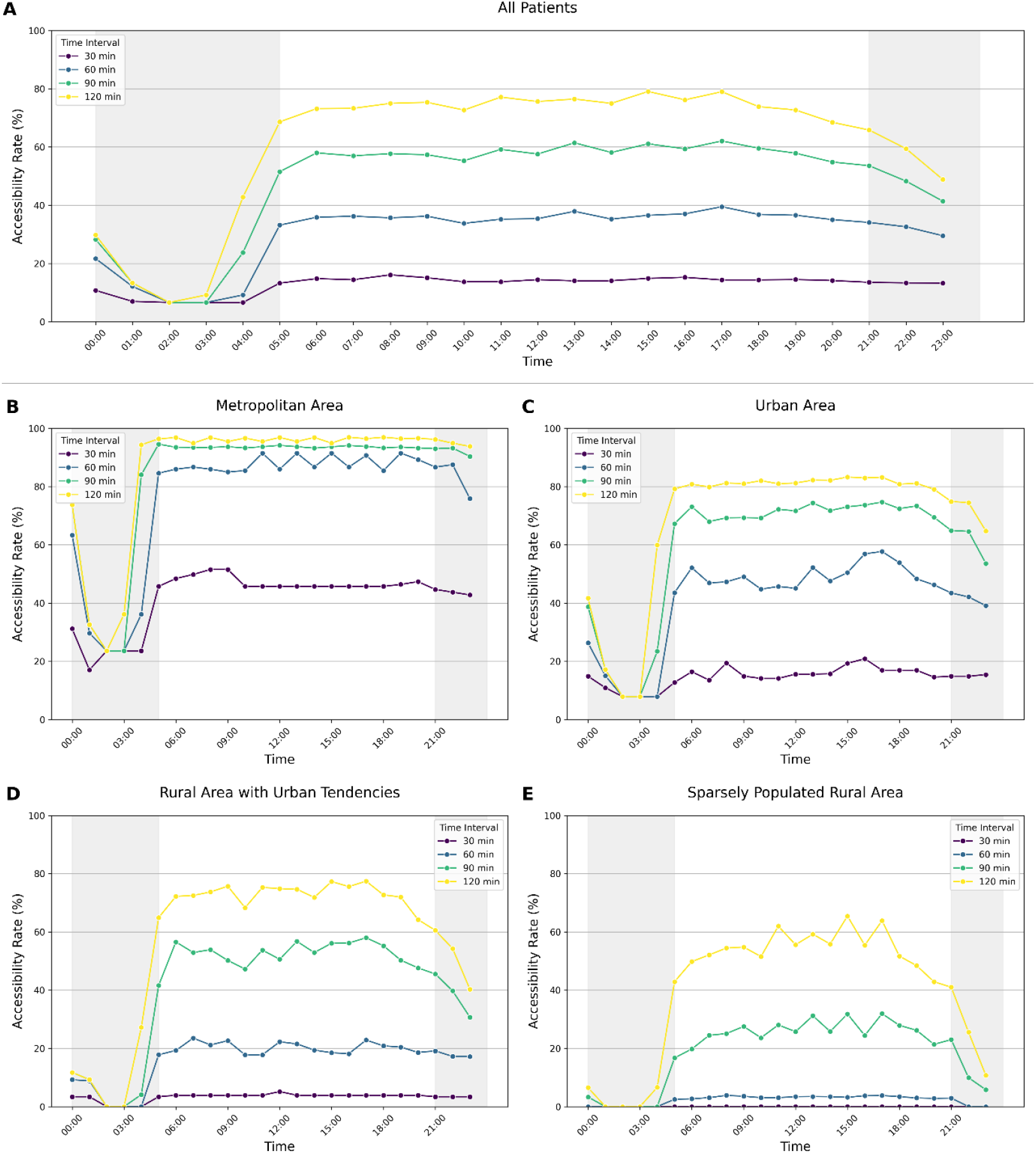
Public transport accessibility for patients from the WERA cohort, depending on time of day and spatial category of residence. (A) Accessibility rates for all patients. (B) Accessibility rates for patients living in metropolitan areas. (C) Accessibility rates for patients living in urban areas. (D) Accessibility rates for patients living in rural areas with urban tendencies. (E) Accessibility rates for patients living in sparsely populated rural areas.

Moreover, we compared public transportation accessibility rates across different spatial categories and found notable differences (Figure 4B–E). During the daytime, approximately 87.8% of patients living in metropolitan areas could reach a WERA site within a maximum travel time of 60 minutes (Figure 4B). However, this patient share dropped to around 49.2% for urban areas (Figure 4C), to 21.4% for rural areas with urban tendencies (Figure 4D), and 3.3% for sparsely populated rural areas (Figure 4E). This decrease in public transportation accessibility in more rural areas was particularly evident during nighttime (between 21:00 and 05:00): While accessibility rates in metropolitan areas maintained at a level of approximately 53% throughout the night (Figure 4B), this rate was markedly lower for patients from urban areas (24%) and rural areas with urban tendencies (9%) (Figure 4C, D). Patients from sparsely populated rural areas suffered the strongest decline, with accessibility rates at the 60-minute interval dropping to 0% between 22:00 and 04:00 (Figure 4E).

### Accessibility by taxi services according to transportation budgets

With no private car on hand and limited public transportation, taxi services remain as an alternative mode of transportation. While the time required for taxi travel is comparable to that of private cars, using taxi services can cause an additional financial burden. Figure 5A illustrates the accessibility zones based on taxi fares for a one-way trip to the nearest WERA site. The analysis indicated that large parts of Bavaria and parts of northern Baden-Württemberg had a sufficient connection within a taxi fare range of €150 (Figure 5A, red circles).

**Figure 5.**
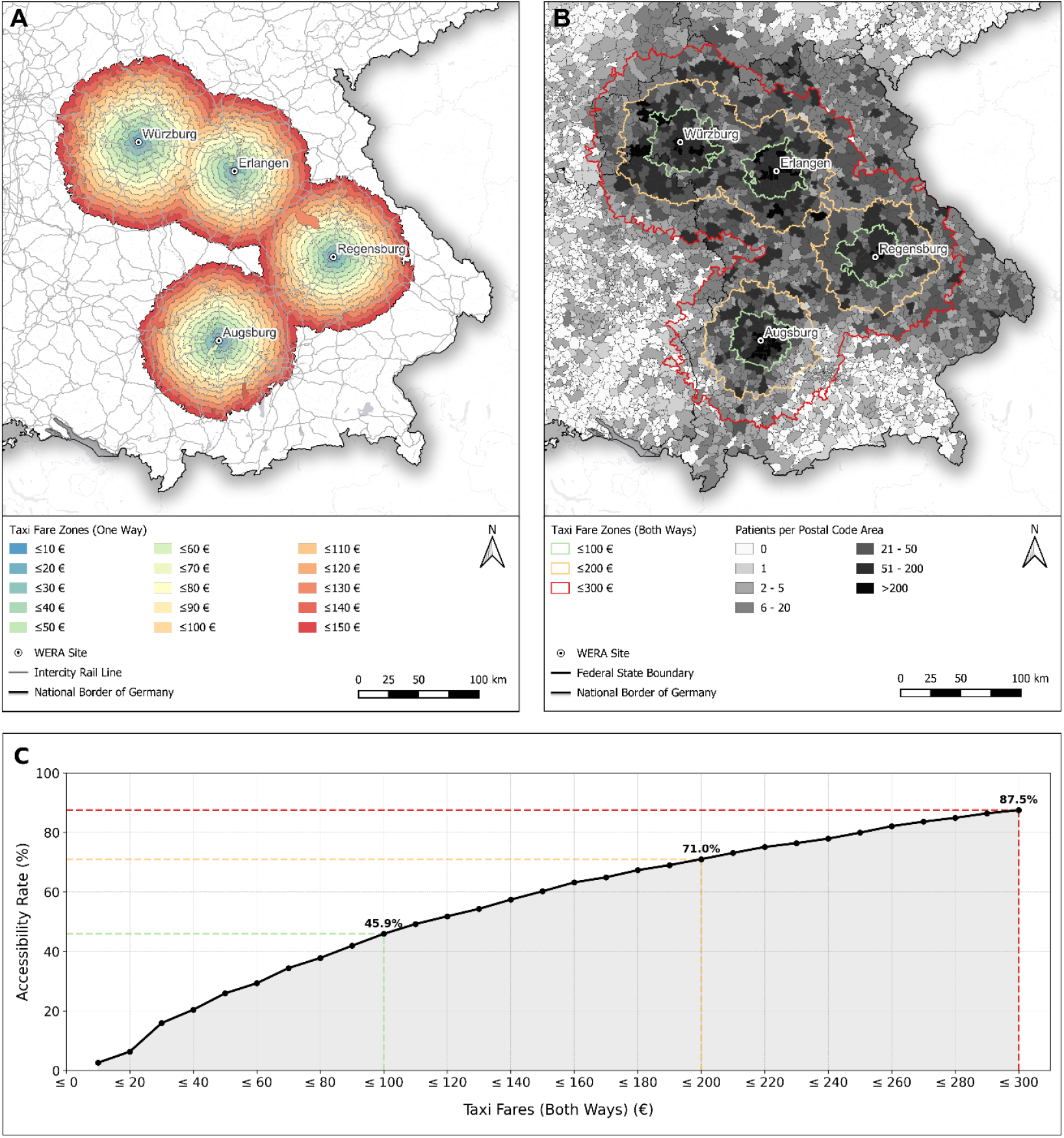
Taxi accessibility for the WERA catchment area. (A) Taxi fare zones starting from €10 to €150 (one-way). (B) Geographical range for virtual taxi budgets of €100, €200, and €300 (two-way). (C) Proportion of WERA cancer patients with cost-neutral access to the closest WERA site based on taxi budgets (fares).

Moreover, we assessed the financial implications of relying on taxi services. We calculated the geographical range for taxi budgets (two-way) between €100 and €300 (Figure 5B). Finally, we calculated the share of WERA patients reaching a cancer center within the given budget. For €100, 45.9% of our patient cohort had access to a WERA site. Increasing the budget to €200 expanded patient access to a share of 71.0%, while a €300 budget ensured that around 87.5% of patients could make a cost-neutral round trip to their closest WERA site.

## DISCUSSION

Given its growing opportunities, we need a better understanding of the factors restricting access to precision oncology. In other words, what keeps cancer patients from receiving innovative cancer care, including molecular tumor boards and biomarker-driven trials? To address this question, we focused on the accessibility of our four WERA cancer centers by different modes of transportation. In a first step, we calculated the accessibility via private car, confirming that our sites could be reached within 120-minute travel time from nearly all places in Bavaria and neighboring regions like northern Baden-Württemberg and Hesse.

### Limited public transportation accessibility and its impact on WERA patients

In sharp contrast to private car use, public transportation accessibility of WERA sites was much lower. Connected areas were primarily located in metropolitan areas, whereas rural areas often had no access within given timeframes. Making things worse, public transportation accessibility strongly depends on the time of day. Patients from regions weakly connected during daytime had no option of public transportation during nighttime. To assess the practical relevance of these findings, we calculated traveling times for a real-world cohort of WERA patients and their places of residence. We found that more than half of our patients lived in rural areas. Especially for patients from these regions, accessibility to the closest WERA site declined sharply after 21:00. Altogether, the limited public transportation accessibility might hinder access to innovative cancer care: on the one hand, patients without a private car might not be able to reach our cancer centers at all. On the other hand, potential patients might not be willing to participate in forms of care like clinical trials if their schedule requires more visits to our cancer centers.

### Relying on taxi services as remaining transportation option

Many patients with frequent visits to our cancer centers rely on taxi services as their regular mode of transportation. Therefore, we calculated the potential financial burden associated with this option. These calculations appear especially relevant for patients in clinical trials: while German health insurance usually covers taxi services for regular hospital visits, this coverage does not apply to trial-related visits. Instead, trial sponsors reimburse taxi costs retrospectively based on a previously negotiated transportation budget. To better understand how this might affect the willingness to participate in a clinical trial, we calculated the “cost-neutral” reach of taxi services based on commonly applied transportation budgets. We found that substantial parts of our patient cohort resided outside the boundaries of these “cost-neutral” zones, potentially indicating further financial toxicities for them.

### Implications for clinicians: “Mind the gap” of public transportation

Our analysis has several limitations. First, we assumed that patients would choose the WERA site located closest to their place of residence. Second, we did not account for multimodal travel patterns involving more than one mode of transportation (e.g., “park and ride” scenarios). Third, our accessibility measures did not account for the use of emergency services. Fourth, even minimal overlaps between isochrones and postal code areas resulted in the entire postal code area being considered capable of reaching a cancer center. Fifth, we calculated travel times assuming optimal traffic conditions, potentially underestimating real-world variability due to congestion or other disruptions. Altogether, these limitations suggest that our approach is rather conservative and may underestimate the true level of travel barriers. Regarding our patient cohort, we cannot rule out that some patients were counted at two WERA sites, e.g., when being hospitalized at one WERA site and receiving a second opinion at another.

However, our findings highlight transportation barriers, which are surely not exclusive to our catchment area.^6–12^ In general, limited accessibility affects the possibility or at least the willingness to receive care from tertiary cancer centers. Therefore, clinicians in these cancer centers need to be aware of accessibility issues and ask their patients for their modes of transportation, especially when they come from regions that are inadequately or not connected by public transportation during nighttime. This patient group may also benefit from digital health solutions and patient-reported outcomes (PROs) such as smartwatch data. By enabling virtual contact with nurses and physicians, these PROs could reduce the number of required visits at distant and “hard to reach” cancer centers.^10,25–26^

In conclusion, understanding local and structural barriers to patient access is key to tailoring effective outreach measures. Therefore, our approach can readily be adopted by other cancer centers and healthcare providers.^27^

## Data Availability

The data used in this study consist of aggregated, anonymized patient counts by postal code area. Due to legal and ethical restrictions, the underlying datasets cannot be shared publicly or upon request.

## CONFLICTS OF INTEREST

The authors declare no conflict of interest.

## ACKNOWLEDGEMENTS

This project is supported by the German Cancer Aid as part of the ONCOnnect Consortium. More information can be found at www.ccc-onconnect.de.

## SUPPLEMENTARY MATERIAL

**Figure S1.**
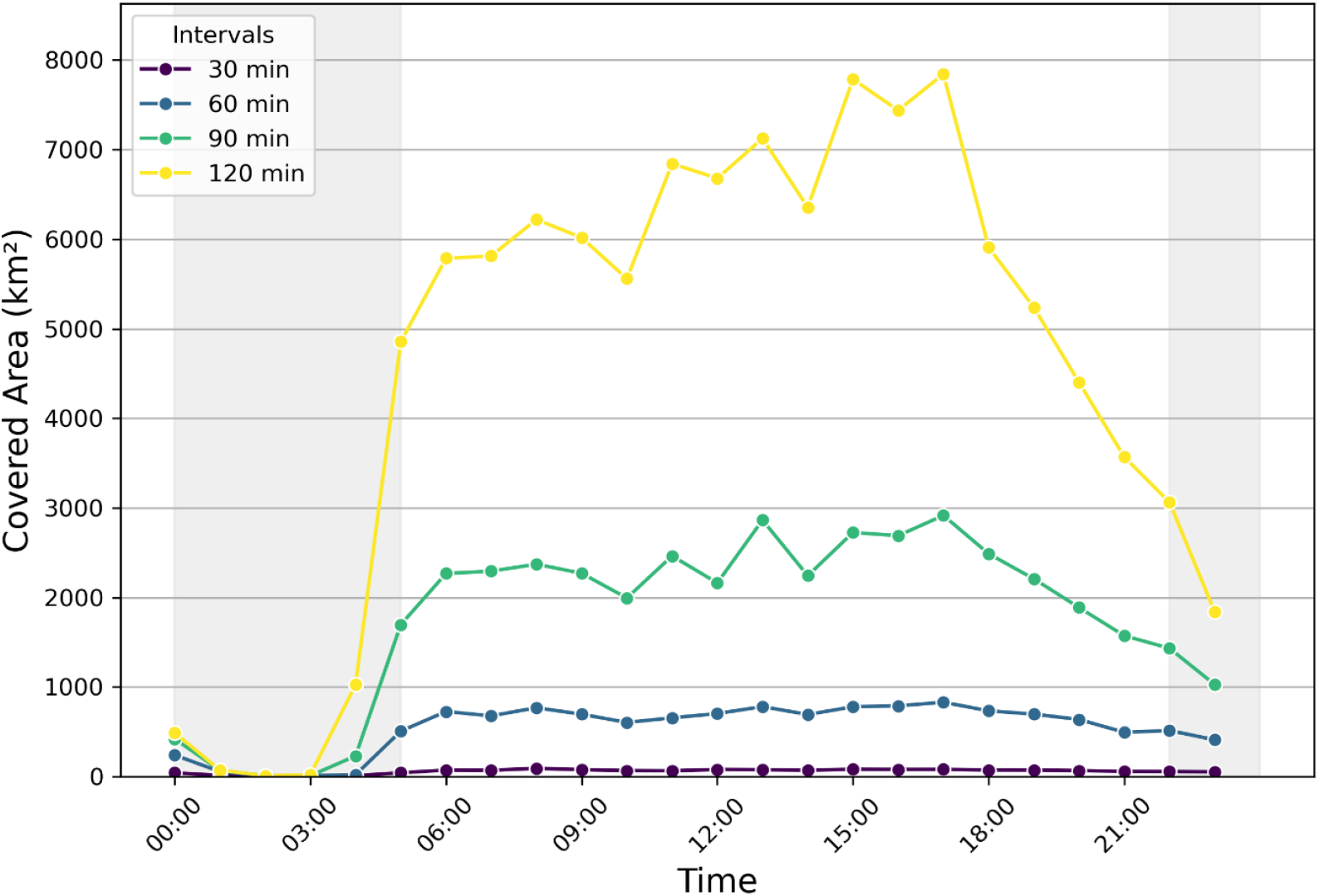
Temporal variation of the covered area by public transportation isochrones around WERA sites. The figure displays the total land area (km^2^) accessible within 30, 60, 90, and 120 minutes, calculated for different times of day. Shaded areas represent night hours with limited public transportation availability.

## Notes

### Competing Interest Statement

The authors have declared no competing interest.

### Author Declarations

Ethics Committee of the University of Regensburg gave ethical approval for this work (25-4415-180).

